# The role of specific and non-specific factors in a brief group psychological intervention for psychological distress: a randomised clinical trial

**DOI:** 10.1101/2024.07.15.24310464

**Authors:** Dharani Keyan, Katie Dawson, Suzanna Azevedo, Srishti Yadav, Jasmine Choi-Christou, Deepthi J. Maliakkal, Mohan K. Pillai, Elizabeth Thomas, Tony S. George, Richard A. Bryant

**Author notes:** Corresponding author: Dharani Keyan, PhD School of Psychology, University of New South Wales, Sydney, Australia.

## Abstract

**Aims:** Although brief psychological interventions in low-and-middle-income-countries have been shown to be effective, they have typically been tested against usual or enhanced usual care (EUC). This design has precluded delineation of the role of specific and non-specific factors in influencing symptom reduction outcomes. This study evaluates the impact of a group psychological intervention (adapted version of WHO’s Problem Management Plus; PM+; titled *Coping with COVID*) against non-directive group Supportive Counselling (SC) on psychological distress during the COVID-19 pandemic in India.

**Methods:** Between October 2020 and December 2022, this two-arm single-blind, controlled trial randomised 183 young adults in Bangalore, India who screened positive for psychological distress to either *Coping with COVID* (n = 91) or SC (n=92), on a 1:1 basis. *Coping with COVID* comprised six weekly small group sessions delivered by videoconferencing that taught stress coping strategies. SC also involved six weekly group sessions that were led by a facilitator and offered non-directive support. The primary outcomes were anxiety and depression as measured by the Hospital Anxiety and Depression Scales (HADS) assessed at baseline, post-intervention, 2-months, and 6-months after treatment. The primary outcome timepoint was the 2-month assessment. Secondary outcomes included generalised worry, positive wellbeing, pandemic-related stress, and suicidal ideation.

**Results:** One hundred and sixty-one participants (88%) were retained at the 2-month follow-up. Intent-to-treat analyses indicated that the *Coping with COVID* condition did not lead to significant reductions in in anxiety (mean difference 0.24 [95% CI, –1.01,1.48], *p*>0.05), or depression (mean difference .03 [95% CI, –1.19, 1.26], *p*>0.05) relative to SC. Similarly, there were no significant differences between conditions for all secondary outcomes.

**Conclusions:** The findings suggest that the benefits of strategies that comprise transdiagnostic scalable psychological interventions may not surpass non-specific factors in driving symptom reduction. There is a need to further evaluate the non-specific factors in scalable psychological programs because focusing on these may have implications for ease of training and implementation.

## Introduction

Scalable interventions for the treatment of common mental disorders (CMDs) in low-and-middle income countries (LMICs) has seen a bourgeoning focus over the past decade (Bryant, 2023). Such evidence-based initiatives have taken a task-sharing framework where non-specialists receive brief trainings to deliver these services across low-resourced settings where skilled mental health practitioners are not readily accessible. Meta-analyses have evidenced that these initiatives are largely beneficial for individuals presenting with CMDs, with small to moderate effects (Singla *et al*., 2017, van Ginneken *et al*., 2013). One of the more widely tested and used interventions is Problem Management Plus (PM+). Developed by the World Health Organization, PM+ is a transdiagnostic intervention aimed to reduce the severity of CMDs in those individuals impacted by adversity (WHO, 2016). Across five sessions, a trained lay provider teaches skills (in either individual or group format) in arousal reduction, management of practical problems, activity engagement, and accessing social support to increase social connection (Dawson *et al*., 2015). The effectiveness of the PM+ program has been validated across diverse populations including migrants, refugee and asylum seekers fleeing in humanitarian crises, and people affected by adversity including conflict and violence (Rahman *et al*., 2016a, Bryant *et al*., 2017b, Rahman, 2019, Bryant, 2022). One meta-analysis found that PM+ administered (in either group or individual format) yielded small to medium effects in reducing the severity of CMDs (Schäfer *et al*., 2023).

The effectiveness of brief psychological interventions in LMICs have typically been tested against usual or enhanced usual care (EUC). This type of control condition refers to the care routinely available to people in their local setting (Gold *et al*., 2017). Given the limited health resources in most LMICs, EUC often might involve little or no mental health care. The use of EUC for the test of new interventions may be justified when the focus is on validating the need for the intervention relative to available resources in an exemplar setting. Tests of interventions against EUC, however, tell us little about the purported mechanisms of action. In the case of PM+, comparisons with EUC preclude delineation between the influence of the content of problem management, arousal reduction, promoting activities, and facilitating social interactions from non-specific factors (e.g., counsellor attention, group support) in contributing to observed gains. There is initial evidence that non-specific factors may account for the effects of scalable intervention. For example, a trial of a WHO self-help program, Self-Help Plus, found that its benefits were no different from a condition that comprised comparable structured activities and supervision (Riello *et al*., 2021). Further, the comparison of brief interventions against EUC potentially may lead to inflated effect sizes because the comparator condition involves a minimal intervention (Mohr *et al*., 2014). It is important to isolate the specific from non-specific benefits of scalable interventions from a public health perspective (Gold *et al*., 2017). The implementation of PM+ can be resource intensive in requiring at least eight days training and ongoing clinical supervision of providers throughout the duration of its implementation (Sijbrandij *et al*., 2017). To justify such resources for a given setting and population, there is an imperative to disentangle the role of such specific and nonspecific factors of scalable interventions such as PM+.

Here we address this issue by reporting a randomised controlled trial of an adapted version of PM+ that was tailored to the psychological needs of people distressed by the COVID-19 pandemic. This program, titled *Coping with COVID*, comprised the same elements as PM+ but was extended to six sessions, and included specific guidance on managing worries during the pandemic (Keyan, 2021). In an initial trial of this program it was shown that it was more effective in reducing anxiety and depression in adults during the pandemic relative to EUC (Bryant *et al*., 2021). In the current trial, the *Coping with COVID* program was delivered by lay peer facilitators to a young adult population during the COVID-19 pandemic in Bangalore, India. Adapted PM+ was compared to non-directive supportive counselling, both of which were delivered over videoconferencing in a group format and led by the same lay peer facilitators. By utilising an active comparator condition, we aimed to assess the potential therapeutic effects of facilitator attention and group support afforded in driving purported effectiveness of the adapted PM+ program. In line with the cumulative findings of the utility of PM+ in reducing the severity of common mental problems across LMICs, it was hypothesised that the adapted PM+ program would lead to greater reductions in common mental health problems relative to non-directive group supportive counselling (SC).

## Methods

### Trial design

This two-arm, single-blind randomised controlled trial was conducted in Bangalore, India in partnership with CHRIST university. Adult participants who screened positive for psychological distress were randomly assigned to either *Coping with COVID* or SC on a 1:1 basis. The primary outcomes were anxiety and depression, and independent assessments were conducted at baseline, post-intervention, 2-months, and 6-months after treatment. The primary outcome timepoint was the 2-month assessment.

The protocol was prospectively registered on Australian New Zealand Clinical Trials Registry (ACTRN12621001064897) and received approval through the CHRIST University Research Ethics Committee (ID: CU: RCEC/64/10/21).

### Participants

The participants were university students studying at CHRIST university who were (a) aged ≥ 18 years, (b) English-speaking with sufficient language comprehension, and (c) scored ≥ 20 on the Kessler Psychological Distress Scale (K10; (Andrews and Slade, 2001)). The K10 is a well-validated 10-item measure of psychological distress, where a cut-off of 20 has been previously used to identify significant distress in adults (Keyan *et al*., 2024). Participants were excluded if they reported: (a) current psychosis, (b) imminent suicidal risk, (c) current substance dependence (but not abuse), or (d) not having access to internet-based videoconferencing. Individuals were recruited through university-wide advertising. Participants provided informed consent for both the screening, and enrolment to the trial.

A simple random assignment was used to allocate participants to either *Coping with COVID* or SC on a 1:1 basis in which randomisation was not stratified on any factor. Randomisation was conducted by staff at UNSW in Sydney who were independent of the trial using computerised software that generated random number sequences. All assessments were completed via online surveys. Access to assessment data was restricted to the research assistant team blinded to intervention allocation throughout the trial.

### Interventions

The *Coping with COVID* program has been detailed in the trial protocol (Keyan *et al*., 2022). Across six weekly group sessions of 60-minute duration (each group comprising 4-5 participants), the facilitator guided participants in psychoeducation relating to common reactions to COVID-19, stress management consisting of breathing retraining, problem management, managing worries during the pandemic, behavioural activation and skills to strengthen social support. For the SC condition, group discussions about how students were coping during the pandemic, ventilation of reactions to common problems experienced and possible solutions were facilitated in a non-directive manner across 6 weekly 60-minute sessions. Both *Coping with COVID* and supportive counselling groups were led by trained male or female peer facilitators who were recruited from CHRIST university campuses across India. These individuals were completing their undergraduate studies in a discipline unrelated to psychology and did not possess prior experience in psychosocial programs. All facilitators received eight days of training in basic counselling skills, delivery of *Coping with COVID* and supportive counselling techniques, group facilitation and self-care. Supervision during practice cycles of *Coping with COVID* and SC was conducted by two clinical psychologists. Practice cycles were used to assess facilitator competency prior to implementation of groups for the trial. To ensure adherence and ongoing support to facilitators, regular weekly supervision was provided remotely by video teleconferencing with a clinical psychologist (DK, KD, SA, or SY) for the duration of the trial.

A trial management committee monitored adverse events and implementation of study procedures. All adverse events were reported by peer facilitators and the local research coordinators to the nominated local clinical psychologist (ET) for further referral to local services.

### Outcomes

All outcome assessments were administered in English. The primary outcome was the severity of depression and anxiety as measured using the Hospital Anxiety and Depression Scales (HADS) (Zigmond and Snaith, 1983), where recommended cut-offs for probable caseness of anxiety and depression is 11 (Stern, 2014). The internal consistency of the HADS in the current sample was robust (0.82) for anxiety and depression. The secondary outcomes assessed the presence of worry symptoms and generalised anxiety with the Generalised Anxiety Disorder scale (GAD-7) where this scale has demonstrated good reliability and validity (Johnson *et al*., 2019) and evidenced higher scores to indicate more severe symptoms (0-21); the COVID Stress Scales (CSS) were used to assess COVID-19 pandemic related stress and anxiety symptoms across five domains including COVID danger and contamination fears, COVID fears about economic consequences, COVID xenophobia, COVID compulsive checking and reassurance seeking, and COVID traumatic stress symptoms (Taylor *et al*., 2020). This scale has evidenced good reliability and internal consistency (Taylor *et al*., 2020). The World Health Organization Well-Being Index (WHO-5) was used assess positive psychological wellbeing as a measure of responsiveness to group both interventions (Krieger *et al*., 2014). The Suicidal Ideation Attributes Scale (SIDAS) was used to both screen the presence of suicidal thoughts and their severity, and as a secondary outcome, where this measure has demonstrated high internal consistency and good internal validity with the GAD-7 and PHQ-9 (van Spijker *et al*., 2014).

### Statistical analyses

Power analyses indicated that an estimated between treatment arm moderate effect size could be achieved at the 2-month primary outcome timepoint through a sample size of approximately100 participants per condition (power of 0.95; alpha = 0.05, two-sided) on the basis of an estimated 10% attrition at the 2-month follow-up time-point.

Pre-planned hypotheses were tested with intent-to-treat analyses. Hierarchical linear models in SPSS (Version 28.) were conducted to study treatment effects because this allows the number of observations to vary between participants and handles missing data by using maximum likelihood estimation methods. Missing data were assumed to be random because after applying a Bonferroni adjustment to accommodate multiple comparisons, participants retained and not retained at 6 months did not differ in terms of baseline characteristics (Table S1). Models included time-of-assessment point, treatment condition, and their interaction. Fixed (intervention, time of assessment) effects and their interactions were entered in unstructured models to yield indices of the relative effects of the treatments; time of assessment included baseline, posttreatment, 2-month, and 6-month follow-up. Fixed effects parameters were tested with the Wald test (t-test, *p* <.05, two-sided) and 95% confidence intervals. Analyses focus on the primary (HADS) and secondary (GAD-7, WHO-5, CSS, and SIDAS scores) outcomes. To determine the robustness of this approach, we repeated the analyses focusing only on participants who completed the 6-month assessment. To determine the relative impacts of the interventions on participants with probable disorder, we repeated the analyses with participants who had probable depression or anxiety at baseline (HADS ≥ 11; (Stern, 2014)).

## Results

### Participant characteristics

Participants were recruited between October 2020 and December 2022 (final 6-month follow-up assessments completed on 3^rd^ July 2023). Participants in the two treatment arms did not differ on baseline characteristics (See Table 1). The mean number of sessions attended did not differ between the *Coping with COVID* (*M*=4.4; *SD* =1.73) and SC (*M*=5.01; *SD*=1.25) conditions. There were 183 participants randomised to either *Coping with COVID* (n = 91) or supportive counselling (n=92). The number of participants enrolled in the trial was marginally less than the projected 200 participants, but enrolment needed to consider local factors that restricted ongoing recruitment. Most participants completed the post intervention assessment (167, 91%), and 2-month assessment (161, 88%). Participants who did (168, 92%) and did not (15, 8%) complete the 6-month assessment did not differ on baseline sociodemographic or psychological variables, with the exception that participants who completed the 6-month assessments were marginally older and marginally reported more COVID stress at baseline than those who did not complete the 6-month assessments (see Table S1). The sample consisted of 121 (66%) individuals with a probable disorder; of this subsample, 110 (60.1%) had probable anxiety and 53 (29%) had depression.

**Table 1.** Participant characteristics.

|  | Total<br>(n = 183) | *Coping with*<br>*COVID* (n=91) | SC<br>(n=92) |
| --- | --- | --- | --- |
| Female, n (%) | 127 (69) | 62 (49) | 65 (51) |
| Age, years (S.D.) | 19.36 | 19.34 | 19.38 |
| Education program, n (%) |  |  |  |
| Bachelors | 166 (91) | 81 (44.27) | 85 (46.4) |
| Masters | 16 (8.7) | 9 (4.9) | 7 (3.8) |
| Baseline HADS: depression | 8.72 | 8.69 | 8.75 |
| Baseline HADS: anxiety | 11.66 | 11.54 | 11.78 |
| Baseline GAD-7: generalised anxiety | 10.01 | 9.80 | 10.21 |
| Baseline WHO-5: wellbeing | 10.04 | 10.20 | 9.89 |
| Baseline CCS: covid stress | 24.19 | 23.51 | 24.87 |
| Baseline SIDAS: suicide | 12.03 | 11.30 | 12.76 |
| Probable Depression | 53 | 28 (53%) | 25 (47%) |
| Probable Anxiety | 110 | 52 (47%) | 58 (53%) |

*Note.* *Coping with COVID*, adapted group Problem Management Plus; SC, Supportive counselling; HADS, Hospital Anxiety Depression Scales (subscale score range: 0-21; higher scores indicate elevated anxiety or depression, where subscale scores $\geq 11$ is considered probable clinical disorder); GAD-7, Generalized Anxiety Disorder Scale (total score range 0-21, where higher scores indicate more severe symptoms); WHO-5, World Health Organization Well-Being Index (total score range: 0-25, where higher scores indicate better wellbeing); CCS, COVID Stress Scales (total score range: 0-144, where higher scores indicate higher covid-19 related distress); SIDAS, Suicidal Ideation Attributes Scale (total score range: 0-50).

### Primary Outcome

The primary and secondary outcomes are presented in Table 2. At the 2-month assessment, participants in the *Coping with COVID* intervention did not differ from those in SC on anxiety (mean difference 0.24 [95% CI, –1.01,1.48], *p*>0.05), or depression (mean difference .03 [95% CI, –1.19, 1.26], *p*>0.05) (table 2, fig 1). The conditions did not differ in depression (mean difference .31 [95% CI, –.78, 1.40], *p*>0.05) or anxiety (mean difference 0.28 [95% CI, –.87, .44], *p*>0.05) severity at post intervention, or at 6-month follow-up (depression, mean difference –0.07 [95% CI, –1.32, 1.18], *p*>0.05); anxiety, mean difference – 0.28 [95% CI, –1.52, .96], *p*>0.05). It is worth noting that both *Coping with COVID* and SC displayed a significant reduction in depression (mean difference 2.38 [95% CI, 1.77, 3.00], *p*<0.001), and anxiety from baseline to the 2-month time point (mean difference 1.76 [95% CI, 1.14, 2.39], *p*<0.001), with a moderate effective size for both outcomes (depression: 0.62; anxiety 0.44).

**Fig 1.**
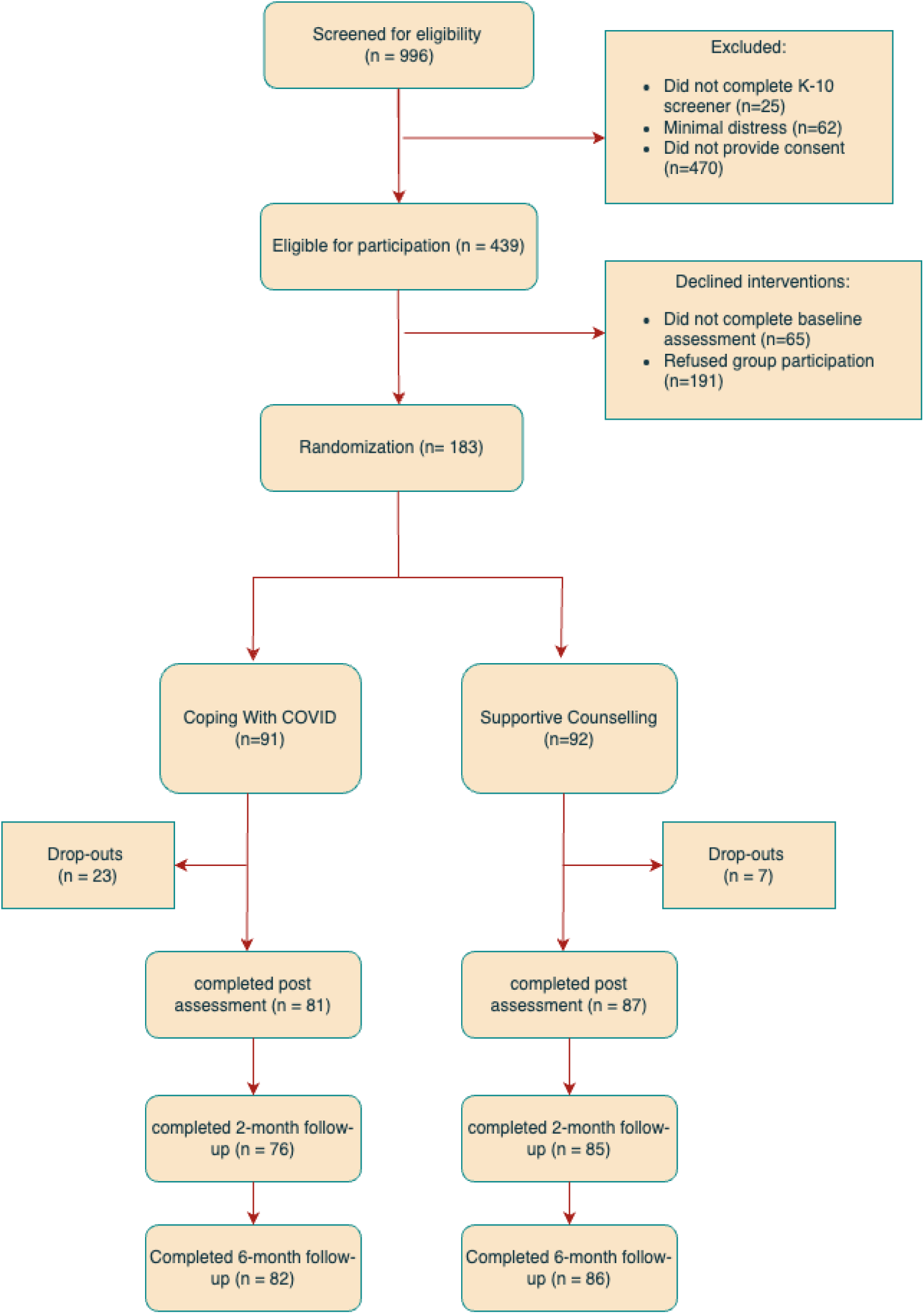
Flow diagram of progress through phases of the randomized trial comparing *Coping with COVID*, the adapted group Problem Management Plus; (PM+) and Supportive counselling in distressed university students in Bangalore, India.

**Table 2.**
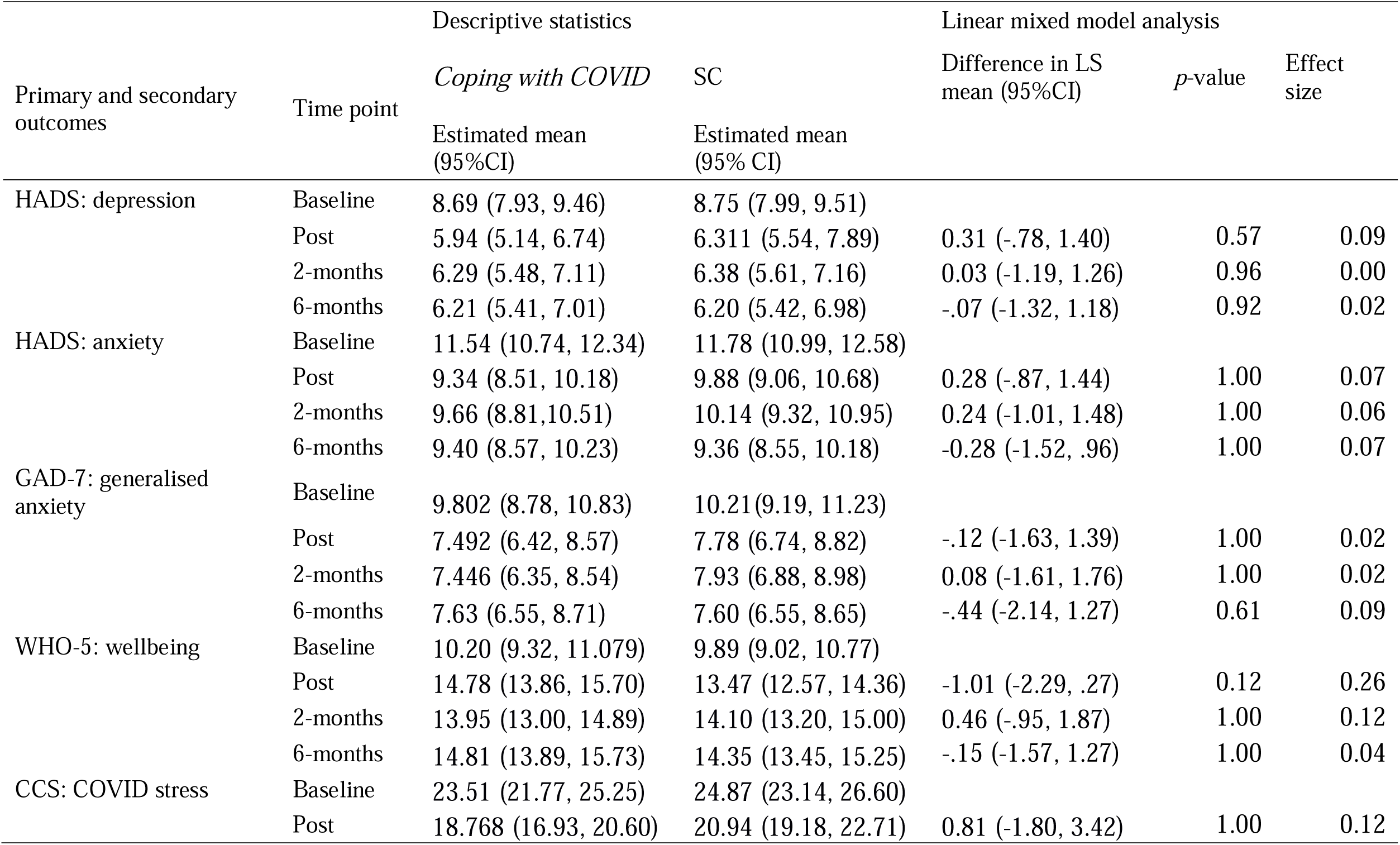
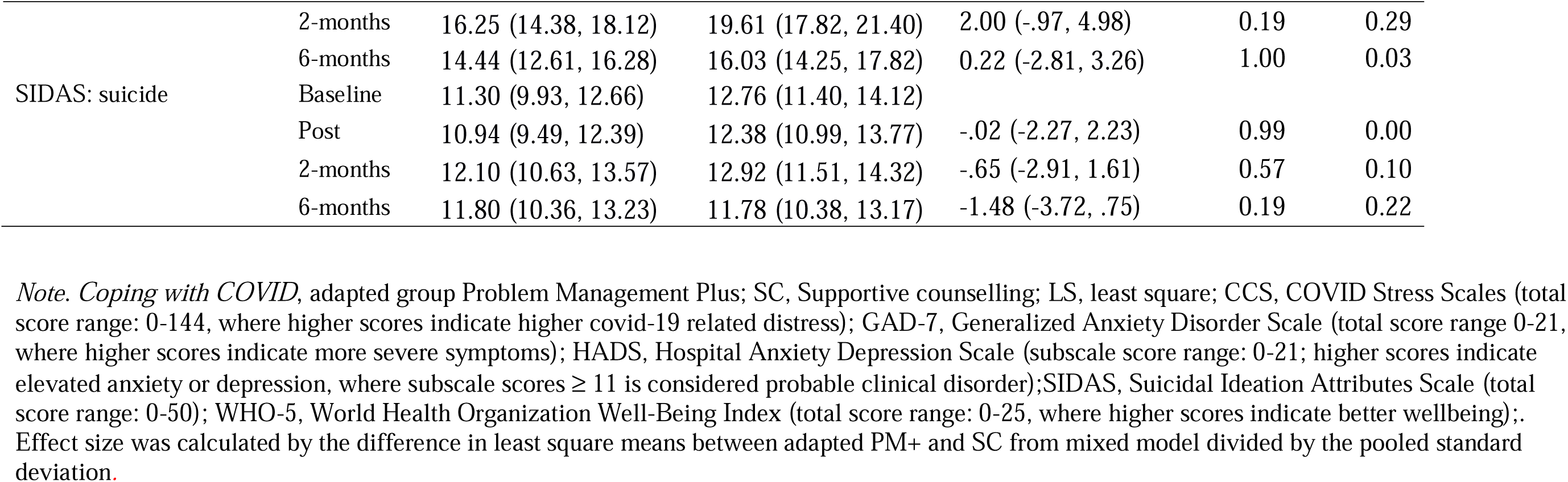
Summary statistics and results from mixed model analysis of primary and secondary outcomes.

### Secondary Outcomes

At the 2-month assessment, there were no differences between conditions on generalised anxiety (mean difference 0.08 [95% CI, –1.61, 1.76], *p*>0.05), wellbeing (mean difference 0.46 [95% CI, –.95, 1.87], *p*>0.05), COVID-related stress (mean difference 2.00 [95% CI, –.97, 4.97], *p*>0.05), or suicidal ideation (mean difference –0.65 [95% CI, –2.91, 1.61], *p*>0.05). Similarly, there were no differences between conditions at post-and 6-month time points (see table 2).

### Secondary Analyses

Repeating the primary analyses with covariates including age and baseline COVID-related stress, similarly, evidenced no differences between *Coping with COVID* and SC on primary or secondary outcomes at all time points (see Table S2). Additionally, participants who were retained at the 6-month follow-up resulted in the same pattern of findings, in that there were no differences between treatment conditions on any primary or secondary outcomes (Table S3). Further, analyses that focussed on participants with probable depression or anxiety similarly reflected the primary analyses (Table S4). That is, *Coping with COVID* did not lead to significantly greater reductions in depression, anxiety, generalised anxiety, COVID-related stress, or suicidal ideation, or increases in wellbeing, relative to SC. No serious adverse events were reported during the trial.

## Discussion

This trial assessed the effectiveness of an adapted PM+ program relative to SC in a distressed young adult population during the COVID-19 pandemic. The major finding was that although both treatment conditions were associated with marked reductions in distress over time, the adapted PM+ program did not demonstrate greater reductions in depression, anxiety, generalised anxiety, COVID-related stress, suicidal ideation, or increased wellbeing relative to SC. This finding does not support our initial hypothesis or accord with previous trials evidencing the effectiveness of PM+ in reducing the severity of common mental health problems (Schäfer *et al*., 2023). This discrepancy between current findings and those of other trials may be attributed to our choice of control condition because no previous trial of PM+ has utilised such an active comparator. The current finding does accord with a previous trial of the scalable Self-Help Plus program which also observed no difference between the treatment and a comparator that controlled for key non-specific factors (Riello *et al*., 2021). The current observation is also consistent with meta-analyses of non-directive counselling that suggest that non-specific elements of treatment can account for a significant proportion of treatment effects in psychotherapy (Cuijpers *et al*., 2012).

The use of non-directive SC as a comparator supports the notion that the choice of control condition is an important consideration that has impacts on outcomes on trials of scalable psychological interventions in LMICs. There are numerous non-specific factors that can influence treatment outcome (for a review, see (Cuijpers, Reijnders and Huibers, 2019). In the context of the adapted PM+ used in the current trial, it is plausible that active listening, encouragement, and group support facilitated reductions in distress. For example, there is abundant evidence that social support can have marked benefits on negative mental health states (Mikulincer, 2016), and so the group format of the intervention may have provided comparable benefits to participants in both treatment conditions. We note that the evidence for non-specific effects of psychotherapeutic interventions is stronger for therapeutic processes rather than clinical outcomes (Priebe *et al*., 2019). Nonetheless, there is evidence that programs that target non-specific factors have been shown to be effective (Priebe *et al*., 2015). Although our current trial does not prove that non-specific factors were causal in the reduction of symptoms, it does suggest that the content of the adapted PM+ program provided no additive benefit over and above the non-specific factors in the SC condition. This pattern points to the need for better understanding of how PM+ strategies may supersede the common non-specific factors on mitigating mental health problems.

We note a number of other potential explanations for the improvement of all participants over time. It is possible that the reductions in distress were a function of contextual factors occurring in India at the time of trial. We note that the stressors associated with the pandemic were frequently changing, and so it is possible that the remission of symptoms in both conditions may have been a function of changing stressors during the pandemic. Alternatively, changes in symptom levels may have been a function of participants acclimatizing to the pandemic, and this may have led to better mental health. Further, it is possible that there was regression to the mean over successive assessments for both conditions. Finally, previous trials of PM+ have been conducted in populations with a history of trauma exposure including conflict, war-affected refugee samples, gender-based violence and other humanitarian crises (Rahman *et al*., 2016b, Bryant *et al*., 2017a). To this end, the current trial may have recruited participants with less severe common mental health problems. This is unlikely given that the proportion of individuals with a probable disorder of depression and anxiety in the current study is comparable to that of previous trials consisting of participants with a history of traumatic experiences (Bryant *et al*., 2022). Despite the potential for these factors to contribute to symptom improvement in both conditions, they do not account for the lack of difference between the active intervention and control conditions.

We note some limitations to this study. First, we did not obtain information relating to mental health services accessed by participants during the trial. Second, a control group that did not receive treatment would have further strengthened our findings relating to comparable effectiveness of adapted PM+ and SC conditions. Third, we did not culturally adapt the primary and secondary outcome measures as participants were English-speaking adults who were completing their university studies at the time of the trial in English. We acknowledge that the lack of proper adaptation of measures may result in failure to capture context-specific idioms and intended constructs most appropriately, however this limitation applies to both treatment arms. Fourth, we acknowledge that attrition at the 6-month follow-up may have been non-random in that those who did not complete these assessments differed in age and baseline pandemic-related COVID stress; however secondary analyses indicate this may not limit the generalisability of our findings. These limitations notwithstanding, strengths of this study include adaptation of PM+ to target pandemic psychological phenomena, use of trained peers as group facilitators, provision of weekly group supervision contact with a clinical psychologist that was matched across conditions, masked online assessments, and very good retention at follow-up.

In conclusion, there are several implications for future research and practice. The current findings suggest that the PM+ strategies may not surpass non-specific factors in driving symptom reduction. The extant literature on PM+ does not help us in disentangling these factors given that all the available trials to date have utilised usual care or EUC as the comparator condition. There is a need to evaluate programs that focus on the non-specific components to determine their efficacy. If it was demonstrated that scalable interventions were primarily beneficial because of non-specific factors, then potential savings could be made in terms of training and scale-up.

## Supplementary material

The supplementary material for this article can be found [doi link]

## Data Availability

The data supporting the findings of our study analyses can be found on the osf data repository.

https://osf.io/5kaf7/?view_only=16f53f28bcd44b77be08c5e9ce83a0d0

## Acknowledgments

The authors acknowledge the dedicated team at CHRIST University, Department of Psychology, who made the preparation and implementation of this trial possible during the pandemic; collegial support and expert input received through the UNSW, Sydney Traumatic Stress Clinic; and the participants, peer facilitators, clinical supervisors, and professionals who provided their time and input into the formative and definitive research guiding this trial.

## Ethical standards

This trial received ethical approval through the CHRIST University Research Ethics Committee (ID: CU: RCEC/64/10/21). The authors assert that all procedures contributing to this work comply with the ethical standards of the relevant institutional committee (as per the CHRIST University Research Ethics Committee regulations).

## Availability of data and materials

The data supporting the findings of our study analyses can be found here: https://osf.io/5kaf7/?view_only=16f53f28bcd44b77be08c5e9ce83a0d0

